# Systematic identification of disease-causing promoter and untranslated region variants in 8,040 undiagnosed individuals with rare disease

**DOI:** 10.1101/2023.09.12.23295416

**Authors:** Alexandra C Martin-Geary, Alexander J M Blakes, Ruebena Dawes, Scott D Findlay, Jenny Lord, Susan Walker, Jonathan Talbot-Martin, Nechama Wieder, Elston N D’Souza, Maria Fernandes, Sarah Hilton, Nayana Lahiri, Christopher Campbell, Sarah Jenkinson, Christian G E L DeGoede, Emily R Anderson, Christopher B. Burge, Stephan J Sanders, Jamie Ellingford, Diana Baralle, Siddharth Banka, Nicola Whiffin

**Author notes:** Correspondence should be addressed to Alexandra Martin-Geary or Nicola Whiffin.

## Abstract

**Background:** Both promoters and untranslated regions (UTRs) have critical regulatory roles, yet variants in these regions are largely excluded from clinical genetic testing due to difficulty in interpreting pathogenicity. The extent to which these regions may harbour diagnoses for individuals with rare disease is currently unknown.

**Methods:** We present a framework for the identification and annotation of potentially deleterious proximal promoter and UTR variants in known dominant disease genes. We use this framework to annotate *de novo* variants (DNVs) in 8,040 undiagnosed individuals in the Genomics England 100,000 genomes project, which were subject to strict region-based filtering, clinical review, and validation studies where possible. In addition, we performed region and variant annotation-based burden testing in 7,862 unrelated probands against matched unaffected controls.

**Results:** We prioritised eleven DNVs and identified an additional variant overlapping one of the eleven. Ten of these twelve variants (82%) are in genes that are a strong match to the individual’s phenotype and six had not previously been identified. Through burden testing, we did not observe a significant enrichment of potentially deleterious promoter and/or UTR variants in individuals with rare disease collectively across any of our region or variant annotations.

**Conclusions:** Overall, we demonstrate the value of screening promoters and UTRs to uncover additional diagnoses for previously undiagnosed individuals with rare disease and provide a framework for doing so without dramatically increasing interpretation burden.

## Background

Current approaches to identify a genetic diagnosis for individuals with rare disease are heavily focussed on protein-coding regions of the genome. Even where genome sequencing data are available, analysis methods often exclude variants that are not in, or immediately adjacent to protein-coding exons. This is in large part due to the difficulty in interpreting variants outside of these regions, but also due to the increased burden of variant review in a clinical context. Studies that have investigated a wider genomic context have successfully identified variants in non-coding regions that cause penetrant Mendelian disease(1–3). The majority of these studies have, however, focussed on small numbers of individuals, specific phenotypes, and/or limited genetic regions. Consequently, we still do not know what proportion of currently genetically undiagnosed individuals with rare disease carry disease-causing variants in non-coding regions.

In this work, we focus on promoters and untranslated regions (UTRs) given that these regions can be confidently linked to known disease genes, and variants within them can significantly disrupt normal gene regulation and have previously been implicated in rare disease(4,5). In short, they provide the best opportunity to expand clinical screening into non-coding regions.

UTRs are regulatory sequences encoded immediately up- and down-stream of the coding sequence (CDS) of protein-coding genes. These regions have important roles in regulating RNA stability, RNA localisation, and the rate of CDS translation(6,7). Variants in UTRs that disrupt these regulatory processes have been shown to cause rare disease through a variety of mechanisms(8). For example, 5’UTR variants that disrupt translation by creating upstream start codons (uAUGs) or perturbing upstream open reading frames (uORFs) cause a range of phenotypes including in genes causing severe developmental disorders (e.g. *NF1*(*9*) and *MEF2C*(*2*), whilst variants directly upstream of the CDS in the *GATA4* gene, that alter the ‘Kozak’ consensus (i.e., the AUG start codon and surrounding motif) have been linked to atrial septal defect(10). Variants resulting in aberrant splicing of the *PAX6* 5’UTR are a frequent cause of aniridia(11). 3’UTR variants that disrupt polyadenylation signals or RNA Binding protein (RBP) binding sites in the α and β-globin genes have been found in individuals with α and β-thalassemia(12). A comprehensive search for 5’UTR variants in retinal disease patients uncovered variants that cause disease through a variety of mechanisms(13).

Proximal promoters comprise an open region of chromatin spanning both up- and down-stream of the transcription start site (TSS) to which transcription factors and polymerase bind to initiate transcription. Variants within promoter regions that disrupt transcription by altering transcription factor binding, or by changing methylation patterns have been identified as being causal of a number of diseases, including *TERT* promoter variants in idiopathic pulmonary fibrosis(14) and *CAMK1D* promoter variants in type 2 diabetes(15). Whilst there are many documented mechanisms through which UTR and promoter variants cause rare phenotypes, our knowledge is likely far from complete. It is also unclear to what extent increasing our understanding of, and regularly including these regions in clinical testing pipelines, will uncover novel diagnoses for currently undiagnosed individuals with rare disorders.

Here, we used the Genomics England 100,000 Genomes Project (GEL) dataset to systematically identify and annotate variants in promoters, UTRs, and UTR introns in 8,040 undiagnosed trios. We developed a reproducible annotation approach with high specificity that can be used in clinical settings without dramatically increasing the number of candidate variants for manual review. After employing strict region-based filters we identified ten likely diagnostic variants, nine *de novo* and one additional overlapping variant. Comparing individuals with rare disorders to matched controls we did not identify a significant burden of rare potentially disruptive variants collectively across any region type or variant annotation, although this may be due to limited statistical power. Our results highlight how promoter and UTR regions can be effectively searched for new diagnoses in rare disease patients and we outline a framework for identification and annotation of such variants in large-scale cohorts.

## Methods

### Identifying known disease genes using PanelApp

PanelApp gene panels were obtained from panelapp.genomicsengland.co.uk using lynx v2.8.9(16), on 12/09/2022. These were filtered to include only the 6,504 genes where the strength of association for one or more gene panel was “green”, corresponding to those with a confident link to the phenotype.

We further filtered to only include genes known to cause a disorder with a dominant mode of inheritance (MOI), inclusive of any genes associated with both dominant and recessive phenotypes. Finally, we selected only genes with transcripts in the MANE v1.0 dataset (17). In total we included 1,536 genes/1,567 transcripts.

### Annotating non-coding regions of interest

Transcripts were defined using MANE v1.0, inclusive of 19,062 MANE ‘select’ and 58 ‘plus clinical’ transcripts(17). UTR exon and intron coordinates were taken directly from the MANE .gff file.

Proximal promoter regions were defined using candidate *cis* regulatory elements (cCREs) obtained from ENCODE(18). Accurate promoter definition is hampered by their tissue specificity. In tissues where a promoter is inactive, it is often marked by a minimal nucleosome free region, but this region may be expanded when the promoter is active. To account for this, as well as promoters that are not annotated at all in the ENCODE dataset, we calculated the average size of all ‘promoter-like’ cCREs that overlap with TSS of MANE transcripts. We calculated the 25th and 75th percentiles of the distribution of distances these cCREs extend up-(25%=181bp; 75%=266bp) and down-stream (25%=67bp; 75%=139bp) of the TSS (Supplementary Figure 1). The 25th percentiles (-181bp to +67bp from TSS) were used to define a ‘minimal’ promoter region.

For transcripts with a cCRE that overlaps the TSS:

● If the cCRE extends ≥ 181bp upstream and ≥ 67bp downstream of the TSS (i.e. at least the minimal 25th percentile definition) the exact region defined by the cCRE is used (Supplementary Figure 1d; n=7,368).
● If the cCRE falls short in either (or both) direction(s), it is extended to reach the 25th percentile distance in that/those direction(s) (Supplementary Figure 1e; extended upstream n=2,953; extended downstream n=2,918, extended in both directions n=464).

For transcripts with no TSS overlapping cCRE (n=5,417), the 75th percentiles are used to define a promoter region that stretches 266bp up- and 139bp down-stream of the TSS.

To ensure identified variants don’t have a protein-coding impact(19) we used bedtools(19) to exclude any positions that intersect with a CDS position in any MANE transcript. In total, we defined 20,417,669 near-coding bases across the 1,567 green PanelApp genes, for an average of 13,030 bases per transcript (min=264, max=791,387), and between 17 and 18,786 per region (Supplementary Table 1). The final set of near-coding regions defined across all green PanelApp genes is in Supplementary Table 2.

### Identifying and filtering *de novo* variants

We used a dataset of previously identified and filtered *de novo* variants (DNVs) within GEL(20), accessed using the RLabKey API(21). We filtered individuals to remove any with subsequently withdrawn consent, and to only include those with a ‘participant type’ of ‘proband’, where neither parent was classified as ‘affected’ or had any associated Human Phenotype Ontology (HPO) terms, and for whom variant calls were on the GRCh38 reference genome. This resulted in an initial set of 9,665 trio probands.

Variants were filtered to only those that passed the most stringent set of GEL filters(22). We removed variants with allele frequency (AF) ≥ 0.00005 and allele count (AC) >=5 in the GEL defined set of 55,603 unrelated individuals or with AF ≥ 0.0005 in any of the major population groups in gnomAD v3.1.1.(23). We restricted our analyses to DNVs within our defined near-coding regions of PanelApp genes with high confidence phenotypic associations (flagged as ‘green’ genes) for the individual’s phenotype. Finally, we excluded participants with an identified coding diagnosis (see below). This resulted in a set of 1,278 DNVs, in 1,094 probands.

### Identifying individuals with existing diagnoses

A list of all participants for whom a confirmed diagnosis was recorded was obtained from the Genomics England ‘exit questionnaire’ table, identified with those for whom the family case was flagged as “solved”. The associated variants were cross referenced with MANE v1.0 coding regions and our promoter, UTR, and UTR intronic regions, with variants mapped onto GRCh38 by the ‘LiftOver’(24) tool, where required.

### Region level variant annotations

Variants for both the *de novo* and burden testing arms of our analysis underwent initial annotation using Ensembl’s variant effect predictor (VEP) v99.1(25) with UTRannotator, SpliceAI v1.3, and CADD v.1.6 plugins(26), as well as custom annotations for PhyloP 100 way vertebrate conservation scores(27), and ClinVar (28) clinical significance annotations (accessed 2022/08/12).

5’UTR variants annotated by UTRannotator(29) were filtered to identify those with the highest likelihood of disrupting translation. To this end, we extracted all variants annotated as:

● uAUG gain resulting in creation of an overlapping open reading frame (oORF) with a strong or moderate Kozak consensus sequence;
● uSTOP loss, with no alternate stop prior to the CDS start (i.e. also resulting in an oORF) and a strong or moderate Kozak consensus sequence;
● uAUG loss, with strong Kozak consensus sequence;
● uFrameshift resulting in an oORF with a strong or moderate Kozak consensus sequence.

For SpliceAI scores, we took the highest delta value across all predictions, with a cutoff of 0.2 for the *de novo* analysis and 0.5 for the burden testing.

Variants across all regions with a ClinVar annotation indicating a benign or protective effect (benign, likely benign, benign/likely benign, protective) were excluded.

Cut-offs for CADD PHRED(26) (25.3) and PhyloP(27) (7.367) scores were taken as the supporting evidence thresholds from Pejaver *et al*(*30*). For variants with multiple recorded scores, the maximum was taken. We note that these scores were calibrated for missense variants, but no alternative exists for non-coding region variants due to the paucity of variants available to benchmark against. Due to this, CADD PHRED and PhyloP scores were not used to annotate variants in deep intronic regions (>20bp from the end of the exon), to reduce noise.

Internal ribosome entry site (IRES) data were obtained from IRESbase(31) on 23/08/2022, microRNA (miRNA) binding sites were obtained from the literature(32–35), and downstream open reading frame (dORF) coordinates were obtained from Chothani *et al*(*36*). For each, locations were cross referenced with our variant positions, and any intersecting variant flagged.

Given the large proportion of variants that fall into miRNA and IRES sites, we excluded any variants that also had a CADD ≤ 22.7 or PhyloP ≤ 1.879. These scores were suggested by Pejaver *et al* as supporting evidence for a benign classification(30).

Kozak consensus sequence variants in the -3 position were identified with reference to MANE v1.0 CDS start positions (i.e. the R of the gccRccAUGG motif). Any variant that changed a reference A or G to a C or T was annotated as potentially Kozak disrupting(37).

RNA binding protein predictions were generated using the methods detailed in Findlay *et al*(*38*) for all possible variants within motifs that are proximal to ENCODE eCLIP sites, that are also high affinity sites as predicted by RBPamp(39). These were intersected with our variants and filtered to retain only those with a reference affinity of ≥ 0.1 and with an impact of ‘loss of binding’ predicted by the RBP binding affinity model (defined as alternative allele affinity / reference allele affinity < ⅓).

Using MANE v1.0 mRNA sequences, we identified the locations of all 3’UTR AAUAAA and AUUAAA polyA signal motifs. We filtered intersecting variants to those that did not result in the creation of an alternative known motif (AAUAAA, AUUAAA, AGUAAA, UAUAAA, CAUAAA, GAUAAA, AAUAUA, AAUACA, AAUAGA, AAAAAG, ACUAAA, or AAAAAA)(40).

Transcription factor binding site (TFBS) locations were obtained from ENCODE(41) and converted using bigBedToBed(24) on the command line, resulting in 4,465,728 TFBS footprints. Any variant not within a footprint identified by ENCODE as falling within the ‘core’ region of a DNase I hypersensitive (DHS) peak was excluded. Remaining variants were annotated using FABIAN(42), limiting to only transcription factor flexible models as these have been shown to outperform positional weight matrices(43). The resultant data was transformed to produce one score per transcription factor, per variant:

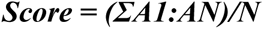

Where A is each model’s predicted change in binding affinity and ‘*N*’ is the total number of these predictions provided for that transcription factor. Scores ≥ 0.04 were recorded as predicted gain and those ≤ -0.04 as predicted loss. For each variant we then calculated the mean gain/loss/total scores and retained any variant with a loss score ≤ -0.4.

### Clinical Review of candidate variants

For each participant carrying a candidate diagnostic *de novo* variant, we compared the similarity between the HPO terms assigned at recruitment with the phenotype expected for a heterozygous loss of function variant in the gene. Variants were interpreted under the assumption that they caused loss-of-function (LoF) and were of high penetrance. Expected phenotypes for each gene were sought from OMIM and the published literature. Where we identified a plausible phenotype match, we raised a clinical collaboration request with Genomics England to confirm or refute our findings by collaboration with the recruiting clinician.

### Defining case and control sets for burden testing

From GEL version 15, we selected participants meeting all of the following criteria:

1. Variants called on genome build ‘GRCh38 and with delivery version ‘V4’
2. Consent not subsequently withdrawn
3. Karyotype one of ‘XX’, ‘XY’, ‘NA’, ‘Other’ and karyotypic and phenotypic sex not in conflict

Cases were defined as:

1. Individuals with a participant type of ‘proband’
2. With at least one ‘green’ PanelApp gene in a virtual gene panel assigned to them
3. Without an existing coding diagnosis (see above).

Controls were taken as the unaffected parents of participants with rare disease. Defined as:

1. Participant type is ‘Mother’ or ‘Father’
2. Affected status is ‘Unaffected’
3. No recorded HPO terms

The genetically-inferred ancestry of each participant, as calculated by GEL, was obtained from LabKey. Participants with a single origin ancestry match of 99% or greater were retained and defined as that ancestry(44). Through this approach, we defined a total of 19,220 cases and 20,683 controls.

### Filtering aggregated variants

Variants within MANEv1.0 transcripts, for all potential case and control participants that passed all internal QC filters were extracted from the aggregated variant VCF files in GEL(45).

In line with recommendations from Pedersen *et al*(*46*) we filtered variants to those with genotype quality (GQ) ≥ 20, read depth (DP) ≥ 10, missingness ≤ 5% heterozygous allele balance (AB) 0.2 ≤ AB ≤ 0.8, and homozygous AB ≤ 0.02. If a variant call failed one or more of these filters in 25% or more cases that call was excluded. We further filtered to only those with GEL internal and gnomAD (v3.1.1; in any population) AF ≤ 0.0001. We retained a total of 18,498,584 variants, a mean variant count per individual of 463.59 (461.74 in cases and 465.74 in controls).

### Participant Matching

To exclude any individuals with very high numbers of called variants (suggestive of systematic error), we calculated a population-specific Z-score per participant as follows:

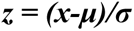

Where ‘*x’* is the variant count in that participant, across all MANE transcripts, from the start of the promoter to the transcript end, ‘*μ* ‘is the population mean, and ‘*σ*’ is the population standard deviation, where the population is all individuals defined of the same genetic ancestry (see above). Participants with a Z-score of ± 2 were dropped (N=1,560, 728 probands, 832 controls) resulting in a set of 18,492 probands and 19,851 control participants.

Within each cohort, we removed individuals with KING score(47) ≥ 0.0442 within the Genomics England relatedness data(44), indicative of being a 3rd degree relative, by randomly selecting one participant for removal in an iterative process until no further relatedness in individuals was detected.

We then matched each proband 1:1 with a single control participant by sex and genetically-inferred ancestry, ensuring that the matched proband and control did not share a family ID. The resultant matched cohort consisted of 18,304 probands, paired with 11,641 unique controls. To avoid potential biases when matching participants caused by low population numbers, we limited to genetically-inferred ancestries where the number of both case and control participants was > 200. This resulted in a cohort of 17,641 case probands, and 11,227 control participants with either European or South Asian genetically-inferred ancestry (Supplementary Table 3). To limit bias due to the presence of large gene panels, this cohort was then limited to probands who had 100 or fewer green dominant genes assigned (Supplementary Figure 2), resulting in a final cohort of 7,862 probands and 6,371 matched controls.

### Burden testing

Aggregated variants filtered as above were further restricted to those with AF ≤ 0.00005 for both internal and gnomAD major population frequencies and to exclude any with an allele count (AC) across the entirety of AggV2 of >= 5. These 1,079,616 variants were annotated and filtered with reference to the annotations described above, with the addition of a more stringent SpliceAI threshold of 0.5 (Supplementary Figure 3).

A simple burden test was performed across all defined near-coding region and variant annotations comparing individuals that had one or more annotated variants meeting our criteria in any near coding region to those that did not, using a Fisher’s exact test performed in R (Supplementary Table 3). The test was repeated for each region annotation separately, using Bonferroni correction for multiple testing.

To estimate the number of participants required to see a significant enrichment across all region and variant annotations, we iteratively increased the number of case and control participants by 1, while maintaining the proportion of observed cases and controls with candidate variants. Fisher’s tests were performed for each iteration, until the resulting *P*-value was ≤0.0031, a Bonferroni adjusted threshold accounting for 16 tests.

### RNA sequencing

Blood was collected from a subset of 100,000 Genomes Project probands in PaxGene tubes to preserve RNA at the time of recruitment. RNA was extracted, depleted of globin and ribosomal RNAs, and subjected to sequencing by Illumina using 100bp paired-end reads, with a mean of 102M mapped reads per individual. Alignment was performed using Illumina’s DRAGEN pipeline. IGV(48) was used to inspect sequencing reads and generate Sashimi plots to show splicing junctions supported by 5 or more reads in areas of interest. FRASER2(49) and OUTRIDER(50) were used to detect abnormal splicing events and expression differences with 499 samples used as controls.

### DNA methylation analyses

DNA methylation array testing was performed on a diagnostic basis as described previously(51,52).

### Availability of data and materials

The datasets supporting the conclusions of this article are available in the near-coding annotation github repository (https://github.com/Computational-Rare-Disease-Genomics-WHG/Near_coding_annotation).

Unless otherwise stated, all analyses were performed using R Statistical software version 4.0.2(53), with the packages ‘dplyr’(54), ‘tidyr’(55), ‘stringr’(56), ‘Rlabkey’(21), ’UpSetR’(57), and ‘ggplot2’(58).

## Results

### Strict region-specific filtering prioritises likely deleterious de novo promoter and UTR variants

We identified 767,063 rare (AF ≤ 0.005%) high-confidence DNVs in 10,665 trio probands in GEL (71.9 per proband), 685,438 of these variants were in participants whose parents were classed as unaffected and had no HPO terms (9,665 probands). Of the remaining probands, 8,040 did not have an existing confirmed diagnosis attributed to a coding variant (576,030 variants, 71.6 per proband). We filtered to include only DNVs in UTR exons and introns (both defined using MANEv1.0 transcripts), and promoter regions (defined by ENCODE candidate *cis* regulatory elements; see Methods). Henceforth, we collectively refer to these regions as ‘near-coding’. We limited our analysis to variants which fell in or near known monogenic disorder genes (3,316 variants) and filtered these to genes which could be plausibly associated with the participant’s phenotype. Accordingly, we filtered for DNVs of genes flagged as ‘green’ in one or more PanelApp(59) gene panel(s) assigned to the individual, and which were associated with disorders with a dominant mode of inheritance. Of note, 309 probands did not have any assigned green dominant genes. In total, we proceeded with 1,311 candidate DNVs in 1,118 probands.

To identify likely disease-causing DNVs we used a region-specific annotation and filtering approach. We prioritised 5’UTR variants that create uAUGs or disrupt uORFs using UTRannotator(29), that overlap *IRES* defined by IRESbase(31), or that lead to disruption of the Kozak consensus sequence(37). 3’UTR variants were prioritised if they disrupt a polyadenylation site or signal sequence, disrupt a miRNA binding site, disrupt an RBP motif, or if they disrupt the start/stop of a dORF with evidence of translation from ribosome profiling (from Chothani *et al*(*36*)). Given the large numbers of variants annotated as within IRES or miRNA binding sites, these variants were further filtered to remove any with CADD (<22.7) or PhyloP (<1.879) scores in support of being benign(30). Across all UTR exons, and in 5’ and 3’ UTR introns, variants with SpliceAI masked delta scores ≥ 0.2 were prioritised. Promoter variants were prioritised if they are predicted to disrupt a transcription factor binding site using FABIAN(42). Finally, across all region annotations, variants with a CADD score ≥ 25.3 and/or a PhyloP score ≥ 7.367 were flagged (thresholds taken from Pejaver *et al*(*30*)). After filtering to only include variants with one or more of these variant annotations, we retained eleven candidate DNVs (0.8% of the initial 1,311 DNVs) each found in a different individual (Figure 1; Table 1).

**Figure 1:**
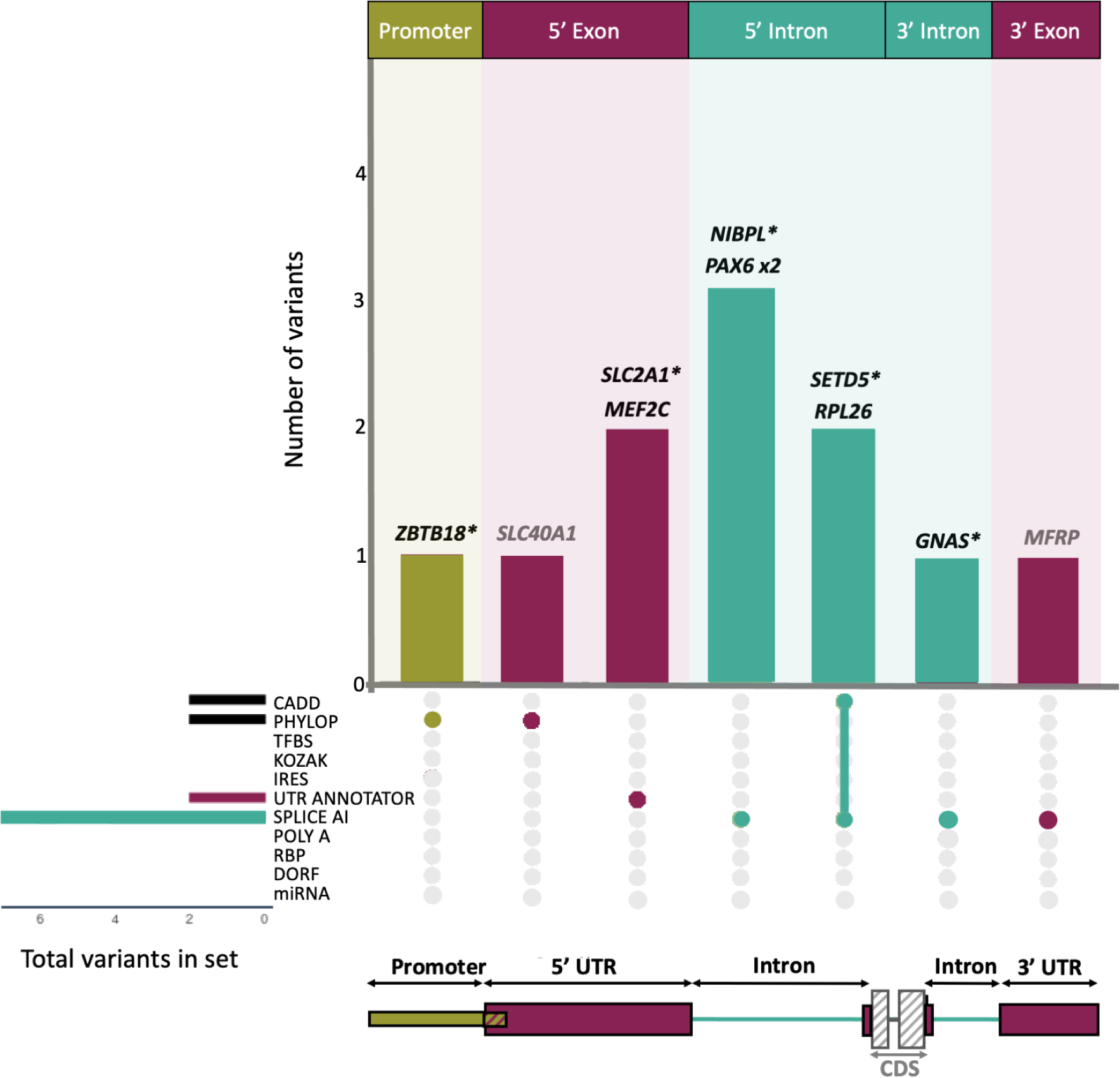
Prioritised *de novo* variants split by region and variant annotations. DNVs were identified from the Genomics England *de novo* dataset in the following regions: Promoter (mustard), UTR exons (raspberry), UTR/Promoter overlapping region (mustard and raspberry stripes), and UTR introns (teal). The gene names corresponding to identified DNVs are written above the corresponding bar. Those in black represent likely diagnoses (nine probands), with those in grey not being a good phenotypic match (two probands). Novel potential diagnoses are marked by an asterisk. Vertical bars in the top panel denote the number of variants identified with specific region and variant annotations that are represented by the bar colour (region annotations), and in the upset plot below (variant annotations). The total number of DNVs with each variant annotation is shown by the horizontal bars to the left of the upset.

**Table 1:** Details of prioritised *de novo* variants. Including the variant annotation which led to it being prioritised, the HPO terms associated with the patient, and whether or not this represents a likely diagnosis. Due to GEL policies, all HPO terms have been re-coded to parent terms with at least one level of abstraction (in some cases up to two) in order to protect the anonymity of participants. Inheritance: autosomal dominant (AD), autosomal recessive (AR), not specified (NS).

| Variant<br>(GRCh38) | Gene (transcript) | Known disease(s) linked to gene | Region<br>annotation | Variant<br>annotation<br>details | Anonymised HPO terms | Possible<br>diagnosis |
| --- | --- | --- | --- | --- | --- | --- |
| Chr1:2440<br>51270<br>C>T | *ZBTB18*<br>(ENST00000358704) | Intellectual Developmental Disorder,<br>(AD; OMIM:612337) | Promoter | PhyloP = 7.426 | Abnormality of higher mental function<br>HP:0011446<br>Abnormality of speech or vocalisation<br>HP:0002167<br>Motor delay HP:0001270 | Yes |
| Chr1:4295<br>8758 C>T | *SLC2A1*<br>(ENST00000426263) | Epilepsy, Idiopathic generalized,<br>susceptibility to, 12, (AD;<br>OMIM:614847),<br>GLUT1 deficiency syndrome 1<br>(AD&AR; OMIM:606777) and 2 | 5'UTR | UTRannotator:<br>uAUG gained.<br>Out of frame<br>oORF<br>(c.-107G>A) | Abnormal nervous system physiology<br>HP:0012638<br>Abnormal homeostasis HP:0012337<br>Phenotypic abnormality HP:0000118<br>Ataxia HP:0001251 | Yes |
|  |  | GLUT1 deficiency syndrome 1, (AD<br>OMIM:612126),<br>Stomatin-deficient chryohydrocytosis<br>with neurologic defects, (AD;<br>OMIM:608885),<br>Dystonia 9, (AD; OMIM: 601042) |  |  | Gait disturbance HP:0001288<br>Neurodevelopmental delay<br>HP:0012758 |  |
| Chr2:1895<br>80685<br>A>C | *SLC40A1*<br>(ENST00000261024) | Hemochromatosis, Type 4, (AD;<br>OMIM:606069) | 5'UTR | PhyloP = 8.189<br>(c.-225T>G) | Azotemia HP:0002157<br>Abnormal hepatic glycogen storage<br>HP:0500030<br>Reduced consciousness/confusion<br>HP:0004372<br>Stroke HP:0001297<br>Language impairment HP:0002463<br>Hypertonia HP:0001276<br>Phenotypic abnormality HP:0000118 | No |
| Chr3:9397<br>978 G>A | *SETD5*<br>(ENST00000402198) | Intellectual developmental disorder,<br>Autosomal Dominant, (AD;<br>OMIM:615761) | 5'UTR<br>Splice | SpliceAI = 0.97<br>Donor loss<br>(c.-177+1G>A) | Decreased body weight HP:0004325<br>Abnormal facial shape HP:0001999<br>Growth delay HP:0001510 | Yes |
|  |  |  |  |  | Aplasia/Hypoplasia of the mandible<br>HP:0009118<br>Intellectual disability HP:0001249<br>Abnormal rib cage morphology<br>HP:0001547<br>Abnormal esophagus physiology<br>HP:0025270<br>Macrocephaly HP:0000256<br>Abdominal symptom HP:0011458<br>Asymmetric growth HP:0100555 |  |
| Chr5:3695<br>3601 T>A | *NIBPL*<br>(ENST00000282516) | Cornelia De Lange syndrome, (AD;<br>OMIM:122470) | 5'UTR<br>Splice | SpliceAI = 0.24<br>Acceptor gain<br>(c.-79-17T>A) | Growth delay HP:0001510<br>Abnormal upper lip morphology<br>HP:0000177<br>Abnormal digit morphology<br>HP:0001167<br>Neurodevelopmental abnormality<br>HP:0012759<br>Decreased head circumference | Yes |
|  |  |  |  |  | HP:0040195 |  |
| Chr5:8882<br>3814 G>A | *MEF2C*<br>(ENST00000504921) | Neurodevelopmental disorder with<br>hypotonia, stereotypic hand<br>movements, and impaired language,<br>(AD; OMIM:613443) | 5'UTR | UTRannotator:<br>uAUG gained.<br>In frame oORF<br>(c.-26C>T) | Abnormality of mouth size<br>HP:0011337<br>Atypical behaviour HP:0000708<br>Neurodevelopmental abnormality<br>HP:0012759<br>Motor delay HP:0001270<br>Reduced visual acuity HP:0007663<br>Decreased body weight HP:0004325<br>Neurodevelopmental delay<br>HP:0012758<br>Aplasia/Hypoplasia of the corpus<br>callosum HP:0007370<br>Abnormal nervous system physiology<br>HP:0012638<br>Abnormal response to social norms<br>HP:5200123<br>Abnormal eyelid morphology | Yes |
|  |  |  |  |  | HP:0000492 |  |
| Chr11:318<br>06844<br>C>T | *PAX6*<br>(ENST00000640368) | Aniridia 1, (AD), OMIM:106210.<br>Foveal Hypoplasia 1, (AD;<br>OMIM:136520),<br>Anterior segment dysgenesis 5, (AD),<br>(AD; OMIM:604229),<br>Keratitis, Hereditary, (AD;<br>OMIM:148190),<br>Coloboma, ocular, autosomal<br>dominant, (AD; OMIM:120200),<br>Coloboma of optic nerve, (AD;<br>OMIM:120430),<br>Optic nerve hypoplasia, bilateral, (AD;<br>OMIM:165550), | 5'UTR<br>Splice | SpliceAI = 0.56<br>Donor loss<br>(c.-52+5G>A) | Phenotypic abnormality HP:0000118<br>Aplasia/Hypoplasia of the iris | Yes |
| Chr11:318<br>06926<br>CT>C | *PAX6*<br>(ENST00000640368) | Aniridia 1, (AD), OMIM:106210.<br>Foveal Hypoplasia 1, (AD;<br>OMIM:136520),<br>Anterior segment dysgenesis 5, (AD), | 5'UTR<br>Splice | SpliceAI = 0.65<br>Acceptor loss<br>(c.-128-2del) | Aplasia/Hypoplasia of the iris | Yes |
|  |  | (AD; OMIM:604229),<br>Keratitis, Hereditary, (AD;<br>OMIM:148190),<br>Coloboma, ocular, autosomal<br>dominant, (AD; OMIM:120200),<br>Coloboma of optic nerve, (AD;<br>OMIM:120430),<br>Optic nerve hypoplasia, bilateral, (AD;<br>OMIM:165550), |  |  |  |  |
| Chr11:119<br>341418<br>C>A | *MFRP*<br>(ENST00000619721) | Microphthalmia, isolated 5, (AR;<br>OMIM:611040)<br>Nanophthalmos 2, (NS; OMIM:<br>609549) | 3'UTR | SpliceAI = 0.56<br>Donor gain<br>(c.*130G>T) | Noncompaction cardiomyopathy<br>Muscular ventricular septal defect<br>HP:0011623<br>Abnormal cardiac septum morphology<br>HP:0001671<br>Neurodevelopmental abnormality<br>HP:0012759<br>Abnormal myocardium morphology<br>HP:0001637 | No |
| Chr17:838<br>2317 T>C | *RPL26*<br>(ENST00000648839) | Diamond-Blackfan anemia 11, (AD;<br>OMIM:614900) | 5'UTR<br>Splice | CADD_Phred =<br>35, SpliceAI =<br>0.97 Acceptor<br>loss<br>(c.-5-2A>G) | Periauricular skin pits HP:0100277<br>Radioulnar synostosis HP:0002974<br>Aplasia/Hypoplasia of the thumb<br>HP:0009601<br>Abnormal zygomatic bone<br>morphology HP:0010668<br>Thickened cortex of bones<br>HP:0100039<br>Phenotypic abnormality HP:0000118<br>Abnormality of the musculoskeletal<br>system HP:0033127<br>Slanting of the palpebral fissure<br>HP:0200006<br>Atrial septal defect HP:0001631 | Yes |
| Chr20:589<br>09654<br>A>G | *GNAS*<br>(ENST00000371075) | Pseudohypoparathyroidism, Type IA,<br>(AD; OMIM:103580) and 1B (AD;<br>OMIM:603233),<br>Pituitary adenoma 3, multiple types, | CDS/3'<br>Intron | SpliceAI = 0.67<br>Acceptor gain<br>(c.*625-30A>G) | Abnormality of the curvature of the<br>cornea HP:0100691<br>Neurodevelopmental delay<br>HP:0012758 | Yes |
|  |  | (NS; OMIM:617686),<br>Pseudopseudohypoparathyroidism,<br>(AD; OMIM:612463),<br>Osseous heteroplasia, progressive,<br>(AD; OMIM:166350),<br>Pseudohypoparathyroidism, Type IC,<br>(AD; OMIM:612462),<br>McCune-Albright syndrome, (AD;<br>OMIM:174800) |  |  | Hypothyroidism HP:0000821<br>Abnormality of body height<br>HP:0000002<br>Phenotypic abnormality HP:0000118<br>Abnormality of refraction HP:0000539<br>Increased body weight HP:0004324 |  |

### Promoter and UTR DNVs provide a diagnosis for undiagnosed individuals with rare disease

Of the eleven remaining candidate variants, nine (82%) were assessed to be a good match for the individual’s phenotype after detailed clinical review (see methods). Three of these had been flagged as diagnostic variants in GEL (in the ‘exit questionnaire’ table) prior to starting this work: two 5’UTR splicing variants in *PAX6* in two individuals with aniridia (OMIM:617141) and one 5’UTR splicing variant in *RPL26* in an individual with a previously undiagnosed monogenic disorder. A further variant, a 5’UTR variant that creates an upstream start codon in *MEF2C*, we previously identified as occurring *de novo* in three unrelated individuals with severe developmental disorders(2). Our approach successfully prioritised all rare DNVs within our candidate regions that had previously been identified as likely diagnostic in GEL. Together, these data demonstrate that our pipeline effectively identifies known diagnostic variants.

Four of the remaining five variants represent likely new diagnoses: (1) a 5’UTR uAUG-creating variant in *SLC2A1* in a patient with GLUT1 deficiency syndrome (OMIM:606777) that was not flagged by GEL as diagnostic, but that has been published previously(3) (Figure 2A). This uAUG is created into a strong start codon context and functional studies support its translation(3). Translation from this uAUG will prevent translation of the downstream CDS, leading to loss-of-function (Figure 2A). After returning this diagnosis to the recruiting clinical team it was classified as Likely Pathogenic and the individual is now on treatment; (2) A *NIPBL* splice disrupting (SpliceAI=0.24) variant 17 bp upstream of the final 5’UTR acceptor site in a participant with a phenotype closely related to Cornelia de Lange syndrome (OMIM:122470, Figure 2B). This variant introduces an AG dinucleotide which is predicted to result in a premature acceptor, however, the positioning of this within the ‘AG exclusion zone’ may also cause skipping of the exon containing the CDS start codon or other splice defects(60) (Figure 2B). The exact impact of this variant will need to be confirmed through RNA studies, but RNA was not available for the patient; (3) A promoter variant that is located in a highly evolutionarily conserved position (PhyloP=7.426) 13 bp upstream of the TSS of *ZBTB18* in a participant with Intellectual disability; (4) A 5’UTR splice-site variant in *SETD5* in an individual originally suspected to have Silver Russell Syndrome (OMIM:180860). This variant is predicted to result in loss of the splice donor (SpliceAI=0.97) of the first 5’UTR exon at the canonical +1 position. DNA methylation signature analysis in this patient revealed an episignature consistent with *SETD5-related neurodevelopmental disorder* (Figure 2C) and no other candidate variants were identified after screening the protein-coding regions of *SETD5*.

**Figure 2:**
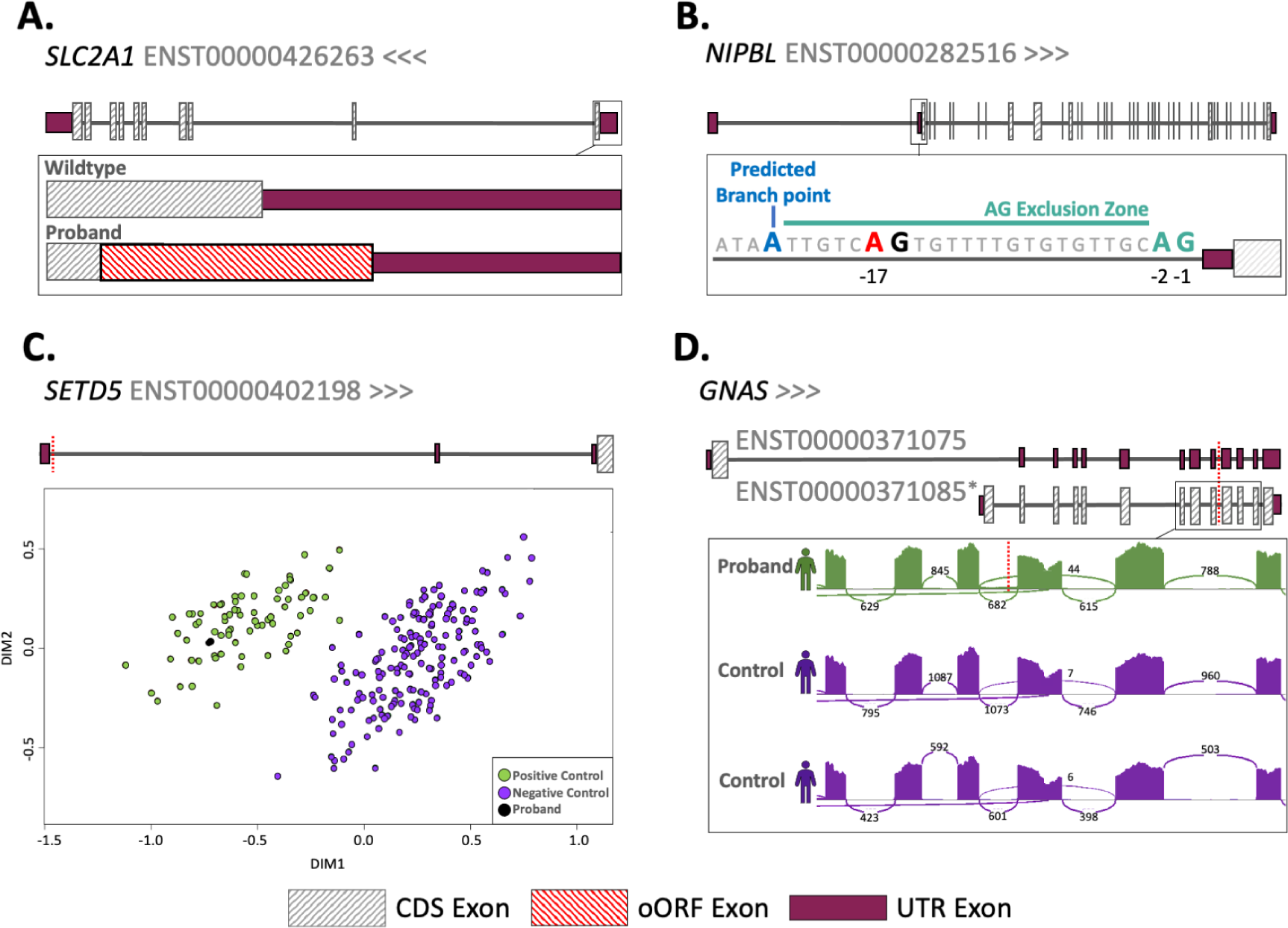
Candidate diagnostic *de novo* variants. A. Gene diagram showing the creation of an out of frame overlapping ORF (oORF; in red) in the *SLC2A1* gene in the proband. B. Illustration of the AG exclusion zone in the *NIPBL* gene. The T>A variant at the -17 position is marked in red, the most strongly predicted branch point (Branchpointer(62) 0.48), directly upstream of the AG exclusion zone is shown in blue. C. Multidimensional scaling plot showing differential methylation in *SETD5.* The position of both variants found in this gene are shown as red dotted lines. D. Sashimi plot showing aberrant splicing in the MANE Plus clinical transcript ENST00000371085. The proband shows some retention of the intron containing the variant (which is marked by a red dotted line) and increased skipping of the following exon compared to the controls (6.06X% vs 0.65X% and 1X%).

We also prioritised a cryptic splice variant in *GNAS* (SpliceAI=0.67) in a participant with hypothyroidism. Whilst we originally annotated this variant as within a 3’UTR intron for the MANE Plus Clinical transcript, the intron is between two CDS exons of the MANE Select transcript. Blood RNA-sequencing from the patient showed evidence of abnormal splicing of the MANE Select transcript, including intron retention (FRASER2 adjusted *P*=1.67×10^-23^), and significantly reduced expression (OUTRIDER adjusted *P*=0.0019; fold-change=0.66; Figure 2D), however the exact mechanism through which this variant could lead to disease is unclear.

For all candidate variants, we checked whether they were found in any other individuals across the full GEL cohort (i.e. not limiting to full trios or DNVs). Whilst we did not observe recurrence of any of the exact variants identified, we did identify a second participant with a different *SETD5* variant at the same genomic position (chr3:9397974 CAAGGT>C, hg38). On closer investigation, this variant is consistent with a germline *de novo*, but it fell just below the required coverage in one parent so it was excluded from the conservative high confidence *de novo* callset. DNA methylation signature analysis also confirmed *SETD5* as the diagnosis in this individual (Figure 2C). In total, we identified a likely disease-causing ‘near-coding’ DNV in ten of 8,040 individuals (0.0012%; nine initially prioritised variants and one additional *SETD5* variant) who did not previously have a coding diagnosis. We classified all six newly identified variants as Likely Pathogenic following the ACMG/AMP guidelines (Supplementary Table 4)(8,61).

### Burden testing does not detect a significant enrichment of variants with any collective region or variant annotation

Given we were able to identify disease-causing near-coding variants using our region-based filtering pipeline, we sought to further use this approach to quantify the enrichment of potentially damaging promoter and UTR variants. However, given the small number of trios within GEL with an unaffected child, and the fact that mutational models to directly assess enrichment of *de novo* variants (by comparing observed to expected numbers) have not been well calibrated for non-coding regions, specifically struggling with the 5’ end of genes(63), we instead used the full aggregated set of inherited and *de novo* variants for our analysis. We matched each of 19,220 rare disease probands without a recorded protein-coding diagnosis, with replacement, to an unrelated unaffected individual from within the rare disease arm (unaffected parents) of GEL as a control on sex and genetically-inferred ancestry (see methods). The control individual was assigned the same dominant, green panelApp genes as had been assigned to the rare disease proband by GEL, allowing us to control for gene and region level differences in mutability (Figure 3). Given the disparity in panel size, with many probands having over 500 assigned green dominant genes (Supplementary Figure 2), to reduce noise we filtered probands to include only those to whom 100 or fewer green dominant PanelApp genes had been assigned (28% of all probands).

**Figure 3:**
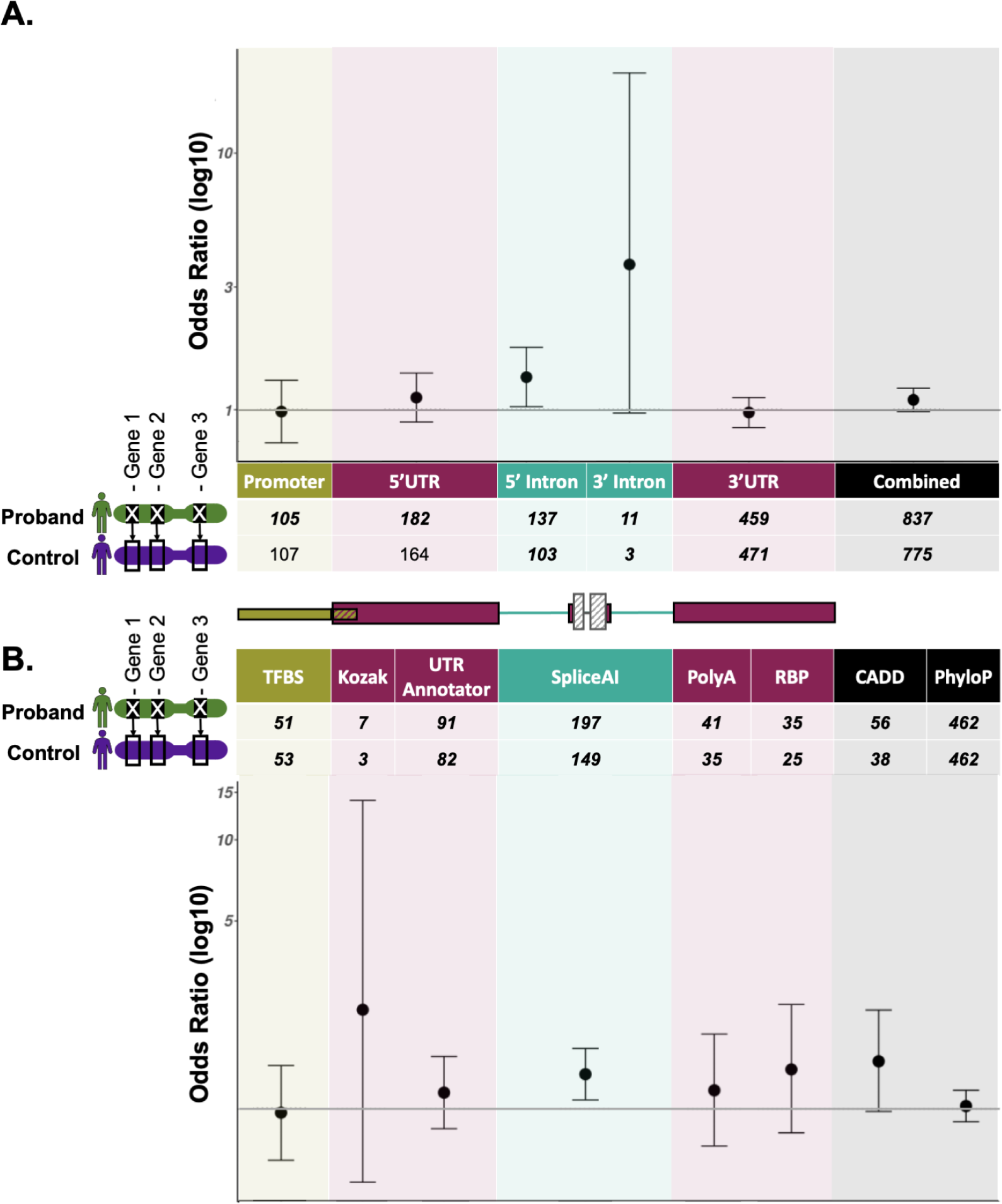
Burden testing results. Counts of variants and odd ratios (log10) testing for an enrichment of variants in cases compared to matched control participants (Fisher’s test), collectively by A. region annotation, and B. variant annotation. Annotation groups with fewer than 10 participants are omitted. Error bars represent 95% confidence intervals. Variants in 5’UTRs (*P*=0.032) and variants with SpliceAI ≥0.5 (*P*=0.008) are enriched in cases over matched controls, but neither is significant after correcting for multiple testing (Bonferroni threshold adjusting for 16 tests =0.0031). Full results are in Supplementary Table 5.

After participant matching, we analysed a final set of 7,862 probands and 6,371 matched controls (1,295 matched controls were partnered with more than one proband). For all individuals, we extracted variants from GEL’s aggregated variant dataset (AggV2) and filtered these using the same region-specific criteria applied to the *de novo* variants. Given that we used a high sensitivity SpliceAI threshold to prioritise DNVs with a high prior of pathogenicity, this was raised to a stricter cutoff of 0.5 for this analysis (Supplementary Figure 3). As we are not powered to analyse individual genes or gene-regions, we performed burden testing collectively across all prioritised variants with the same regional (e.g., 5’UTR) or variant-level (e.g. SpliceAI) annotations, across all participants and their assigned green genes. Whilst we observed a greater number of probands with prioritised variants compared with matched controls for the majority of regional and variant-level annotations we identified a greater number of probands with prioritised variants compared with matched controls, no specific annotation was significantly enriched for variants in cases after correcting for multiple testing (Figure 3; Supplementary Table 5). We also did not observe a significant enrichment when combining across all regions and variant annotations (Fisher’s *P*=0.109*, OR=*1.09, 95% *CI*=[0.981,1.210]).

Assuming a constant ratio between case and control variants, and hence ORs, we would need an estimated 11,610 cases and controls for significance at *P*<0.05 in the combined test across all region and variant annotations, and 26,066 for significance at a study-wide *P*-value threshold of <0.0031, correcting for 16 tests (Supplementary Figure 4).

## Discussion

Here, we have described a framework for the identification and annotation of potentially disease-causing UTR and promoter variants in individuals with rare disease. We show the utility of the approach through identification of ten likely diagnoses in the GEL rare disease cohort. These comprised: three new confirmed diagnoses (*SLC2A1* and 2x *SETD5*) and three new likely diagnoses (*GNAS, NIPBL, ZBTB18*) alongside four previously confirmed diagnostic variants (2x *PAX6*, *RPL26*, and *MEF2C*). This illustrates the importance of expanding diagnostic screening into near coding regions of known disease genes.

In our analysis, we concentrated on variants within or directly adjacent to UTR exons and proximal promoter sequences for three key reasons: (1) the functional link between these regions and the impacted gene is relatively clear; (2) the importance of these regions in gene regulation means that variants within them can have a large impact, even causing complete loss-of-function; and (3) known functional elements within these regions enable us to predict some variant effects. Many of these criteria do not apply to more distal non-coding elements, such as enhancers, which also suffer from redundancy, meaning small variants in any one enhancer may often be unlikely to have a large impact on gene expression and hence disease(64), although there are exceptions(65). Recent work has, however, shown that variants impacting tissue-specific silencer elements may be a frequent cause of some disorders, indicating that these specific elements may have lower levels of redundancy(66,67). More research is needed to clarify the contribution of other non-coding elements to rare monogenic disorders.

A key barrier to routine identification of non-coding variants in clinical settings is the potential dramatic increase in interpretation burden. Here, we employed strict filters based on known regulatory mechanisms, aiming for high specificity. Consequently, a very large proportion of the shortlisted variants (∼82%) were flagged as good diagnostic candidates following clinical review. This illustrates the validity of our method as a highly specific route to finding new diagnoses without dramatically increasing the number of variants that need to be manually reviewed. Here, we focus on *de novo* variants given their high prior probability of being pathogenic. Currently, *de novo* inheritance pattern, clinical fit, and functional validation are essential to identifying and classifying non-coding variants as (likely) pathogenic. Hence it is much harder to identify disease-causing non-coding variants in more heterogeneous conditions and/or disorders where *de novo* variants are not a frequent disease mechanism. However, the same annotation approach can be applied to inherited variants(68).

Despite our strict filtering approach, the relatively modest number of new diagnoses given the size of the GEL cohort suggests that the proportion of currently undiagnosed individuals that will likely be diagnosed through regular assessment of proximal promoter and UTR regions will also be modest. This is in-line with the conclusions of our recent work looking for non-coding variants in recessive disease genes(68). Nevertheless, our diagnostic yield is likely an underestimate. First, we limited our analyses to only genes within a diagnostic panel applied to each individual and, within this we focussed on genes with a clear dominant disease mechanism. Gene agnostic approaches may have greater sensitivity for new diagnoses and allow the identification of candidate novel disease genes. Our study was also limited to MANE transcripts and may miss important variants impacting alternate transcripts. Our strict filtering approach was necessitated by our limited understanding of the ‘regulatory genetic code’, and the paucity of tools to accurately determine non-coding variant deleteriousness and also likely excluded some important variants. Finally, we only removed individuals flagged as ‘solved’ in the GEL ‘exit questionnaire’ as having an existing diagnosis. Many more individuals may have subsequently had likely diagnostic variants returned that were not reflected in the exit questionnaire at the time of analysis, due to ongoing analyses of the cohort.

Amongst our novel diagnoses was a 5’UTR uAUG-creating variant in *SLC2A1*, variants in which cause GLUT1 deficiency syndrome, which is treatable through diet. Hence, our diagnosis changed the clinical management of this patient. The exact same variant was found in a patient with a similar phenotype in 2017(3), the same year the patient was recruited to GEL, but, whilst the variant was deposited in the more specialist Leiden Open Variation Database(69) (I.D: SLC2A1_000036) it did not appear in the more widely used ClinVar database until 2022 (ID:1491299). This highlights the necessity of data sharing through variant databases and the use of these datasets for re-analysis to reduce the lengthy diagnostic odysseys so often faced by individuals with rare disease.

Whilst we expected the excess burden of near-coding variants in cases to be relatively low, our approach was imperfect. In particular, analysing all variants identified in each individual (i.e., inherited as well as *de novo*) across large gene panels likely added a lot of noise. A better approach to assess this enrichment would be using only *de novo* variants, however, the number of trios within GEL where the child is unaffected is very small, and we and others have struggled to correctly optimise mutational models for application at the 5’ end of genes(63). Multiple additional factors also likely contribute to our observed lack of signal. Firstly, we used unaffected parents of rare disease probands as a control and these individuals may be more likely to carry damaging variants (for example variants with reduced penetrance, or variants that modify coding variant penetrance). Secondly, the sizes of gene panels varied substantially between participants, with some containing vast numbers of genes (Supplementary Figure 2). These larger panels likely contribute an overrepresentation of variants that are unlikely to be causal.

## Conclusions

Our understanding of the mechanisms that underlie variation in the non-coding genome is far from complete. Despite this, routine interrogation of these regions with existing knowledge and tools can return valuable genetic diagnoses to patients. Identifying more disease-causing variants in non-coding regions and understanding how they lead to disease will, in turn, increase our understanding of regulatory biology, and enable us to create better tools to identify and annotate these variants. Here, focussing specifically on proximal promoters, UTRs, and UTR introns, we developed a flexible approach for variant annotation and filtering which can be extended and adapted to incorporate new functional variant classes as our understanding of non-coding genome biology increases. Our framework provides a foundation for the systematic analysis of variants in these regions, which can be readily applied to cohorts, and in clinical settings, globally.

## Data Availability

All publicly accessible data produced are available online in the near_coding_annotation GitHub repository.
Any additional data is located in the National Genomic Research Library, which is managed by Genomics England Limited (a wholly owned company of the Department of Health and Social Care).

https://github.com/Computational-Rare-Disease-Genomics-WHG/Near_coding_annotation

## Acknowledgements

NWhiffin and ACM-G are supported by a Sir Henry Dale Fellowship awarded to NWhiffin, jointly funded by the Wellcome Trust and the Royal Society (220134/Z/20/Z). NW, END, and MF are supported by research grant funding from the Rosetrees Trust (PGL19-2/10025). AJMB is supported by a Wellcome PhD Training Fellowship for Clinicians and the 4Ward North PhD Programme for Health Professionals (223521/Z/21/Z). JL is supported by a University of Southampton Anniversary Fellowship. DB is supported by a National Institute for Health Research (NIHR) (RP-2016-07-011) research professorship. SJS is supported by grant funding from the National Institutes of Health (NIH) (R01 MH116999 S.J.S. and U01 MH122681)

This research was made possible through access to data in the National Genomic Research Library, which is managed by Genomics England Limited (a wholly owned company of the Department of Health and Social Care). The National Genomic Research Library holds data provided by patients and collected by the NHS as part of their care and data collected as part of their participation in research. The National Genomic Research Library is funded by the National Institute for Health Research and NHS England. The Wellcome Trust, Cancer Research UK and the Medical Research Council have also funded research infrastructure.

We would like to acknowledge the Genomics England service desk, in particular Daniel Rhodes, Roel Bevers, and Peter O’Donovan, for the support and guidance they provided to make this work possible. We would also like to thank Ana Lisa Taylor Tavares for her assistance in orchestrating connections with clinical collaborators, and both Esther Ng and Matteo Ferla for valuable discussions, and insight.

We acknowledge Episign® for the diagnostic DNA methylation array testing, and are grateful for the support of the NIHR Manchester Biomedical Research Centre (NIHR203308).

