## Supplementary_figures_and_tables for "Systematic identification of disease-causing promoter and untranslated region variants in 8,040 undiagnosed individuals with rare disease"

### Supplementary section

#### Supplementary Tables

**Supplementary Table 1**: Total, and mean size of near-coding region annotations by type.

| Region | Total (bp) | Mean per gene (bp) |
|---|---:|---:|
| Promoter | 543,422 | 347.9 |
| 5'UTR | 372,417 | 239.2 |
| 5'UTR intron | 15,795,406 | 10080.0 |
| 3'UTR | 3,437,362 | 2194.7 |
| 3'UTR intron | 263,304 | 168.0 |
| All Regions | 20,409,032 | 13029.8 |

**Supplementary Table 2**: Genomic coordinates of analysed near-coding regions across green dominant PanelApp genes.

Hosted on GitHub due to size:

https://github.com/Computational-Rare-Disease-Genomics-WHG/Near_coding_annotation

**Supplementary Table 3**: Total and mean variants across promoters and full MANE transcript length, in case and control participants in matched pairs, by genetically inferred ancestry group, and sex (AF 0.0001).

| Sex | Genetically inferred ancestry | Case participants | Control participants | Mean Case variants | Mean Control variants |
|---|---|---|---|---|---|
| Female | European | 7,827 | 5,198 | 430.38 | 428.04 |
| Male | European | 8,096 | 4,881 | 430.47 | 427.91 |
| Female | South Asian | 729 | 538 | 619.10 | 629.93 |
| Male | South Asian | 989 | 610 | 629.94 | 631.29 |

**Supplementary Table 4**: Classifications for the six newly identified DNVs according to the ACMG/AMP guidelines and adaptations for variants in non-coding regions(8,61). Thresholds for PP3 were taken from Pejaver *et al*. 2022 (doi: 10.1016/j.ajhg.2022.10.013) for PhyloP and CADD, and Walker *et al*. 2023 (doi: 10.1101/2023.02.24.23286431) for SpliceAI. *Functional data from Willemsen *et al*. 2017 (doi: 10.1038/ejhg.2017.45).

| Variant | Gene | Applied rule codes | Classification |
|---|---|---|---|
| chr1:g.244051270 C>T | *ZBTB18* | PS2 (de novo); PP3_supporting (PhyloP score); PM2_supporting (absent from gnomADv3.1.2) | Likely Pathogenic |

| | | | |
|---|---|---|---|
| chr1:g.42958758 C>T | *SLC2A1* | PS2 (de novo); PP3_supporting (UTRannotator prediction); PM2_supporting (absent from gnomADv3.1.2); PS3_supporting (published functional studies*) | Likely Pathogenic |
| chr3:g.9397978 G>A | *SETD5* | PS2 (de novo); PP3_moderate (SpliceAI score); PM2_supporting (absent from gnomADv3.1.2); PP4_supporting (methylation studies confirm *SETD5*); PS1_supporting (same nucleotide) | Likely Pathogenic |
| chr3:g.9397974 CAAGGT>C | *SETD5* | PS2 (de novo); PP3_moderate (SpliceAI score); PM2_supporting (absent from gnomADv3.1.2); PP4_supporting (methylation studies confirm *SETD5*); PS1_supporting (same nucleotide) | Likely Pathogenic |
| chr5:g.36953601 T>A | *NIBPL* | PS2 (de novo); PP3_moderate (SpliceAI score); PM2_supporting (absent from gnomADv3.1.2) | Likely Pathogenic |
| chr20:g.58909654 A>G | *GNAS* | PS2 (de novo); PM2_supporting (absent from gnomADv3.1.2); PS3_supporting (evidence of aberant splicing and significant reduction in expression compared to 499 controls shown in RNA-seq) | Likely Pathogenic |

**Supplementary Table 5**: Burden testing of prioritised variants in case and control participants by region and variant annotation. The proportion of total participants (N=7,862) are shown in the control, and case participant columns in round brackets, and 95% confidence intervals are shown in square brackets alongside the odds ratios.

| Region | Control Participants | Case Participants | Fisher's *P*-value | Odds Ratio |
|---|---|---|---|---|
| ALL | 775 (9.86%) | 837 (10.65%) | 0.109 | 1.090 [0.981,1.210] |
| Promoter | 107 (1.36%) | 105 (1.34%) | 0.945 | 0.981 [0.741,1.299] |
| 5'UTR | 164 (2.09%) | 182 (2.31%) | 0.356 | 1.112 [0.894,1.386] |
| 5'UTR intron | 103 (1.31%) | 137 (1.74%) | 0.032 | 1.336 [1.025,1.746] |
| 3'UTR | 471 (5.99%) | 459 (5.84%) | 0.710 | 0.973 [0.850,1.113] |
| 3'UTR intron | 3 (0.04%) | 11 (0.14%) | 0.057 | 3.670 [0.969,20.494] |
| **Annotation** | | | | |
| CADD | 38 (0.48%) | 57 (0.73%) | 0.063 | 1.504 [0.979,2.333] |
| PhyloP | 462 (5.88%) | 473 (6.02%) | 0.736 | 1.025 [0.896,1.173] |
| TFBS | 53 (0.67%) | 51 (0.65%) | 0.922 | 0.962 [0.641,1.442] |
| KOZAK | 3 (0.04%) | 7 (0.09%) | 0.344 | 2.335 [0.533,13.995] |
| IRES | 0 (0.00%) | 1 (0.01%) | | |
| UTR Annotator | 82 (1.04%) | 94 (1.20%) | 0.404 | 1.148 [0.843,1.566] |
| SpliceAI | 149 (1.90%) | 199 (2.53%) | 0.008 | 1.344 [1.079,1.678] |
| PolyA | 35 (0.45%) | 41 (0.52%) | 0.566 | 1.172 [0.728,1.898] |
| RBP | 25 (0.32%) | 35 (0.45%) | 0.244 | 1.402 [0.815,2.446] |

| | | |
|---|---|---|
| dORF | 7 (0.09%) | 2 (0.03%) |

#### Supplementary Figures

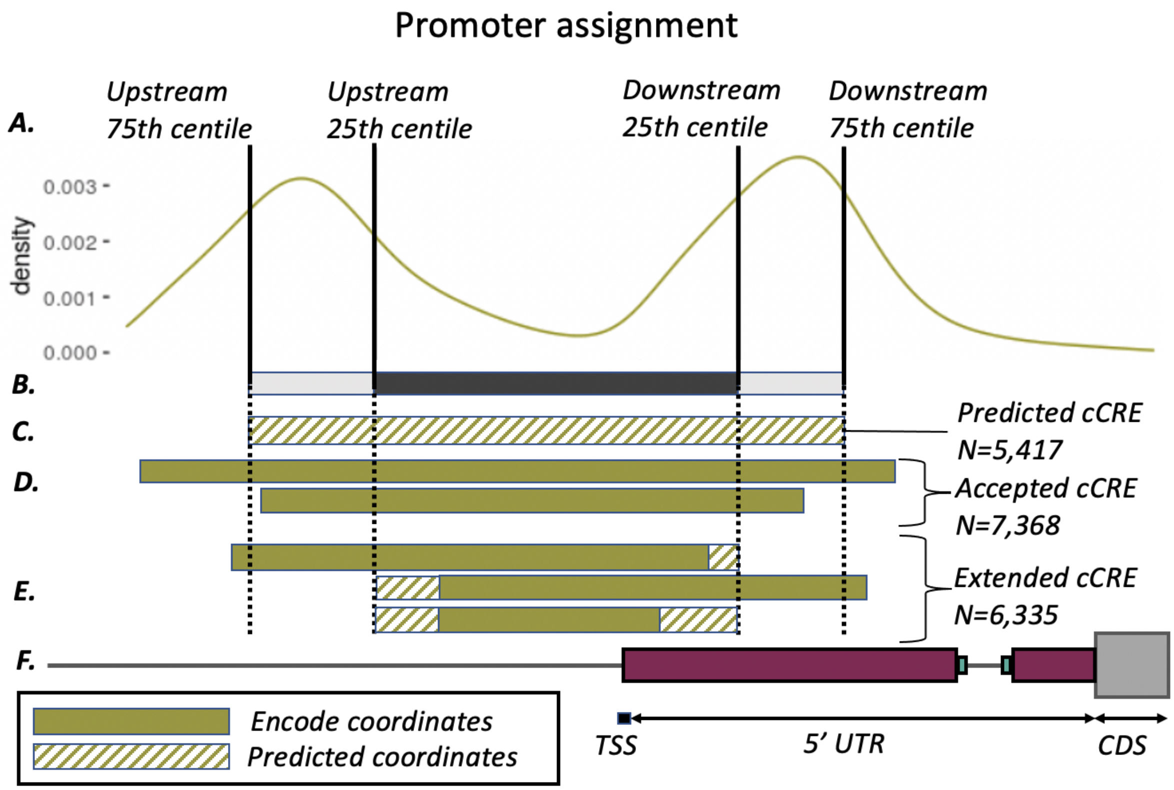

**Supplementary Figure 1**: Graphical representation of promoter assignment with reference to the TSS (F). For all cCREs that overlapped a TSS the total distance the cCRE extends in each direction (upstream/5' and downstream/3') was calculated (A). The 25th and 75th percentiles of the distance in each direction was calculated to form the basis of a 'minimal' (B, dark grey) and 'maximal' (B, light grey) promoter. For genes without an overlapping cCRE a promoter was predicted using the 'maximal' promoter criteria (C, shaded line). For genes with an overlapping cCRE that extended beyond the 25th centile in both directions (D) the cCRE was accepted without modification. For genes where the overlapping cCRE

(E, solid line) fell short of the 25th centile in either direction, this was extended to match the coordinates of the 'minimal' promoter (E, shaded line).

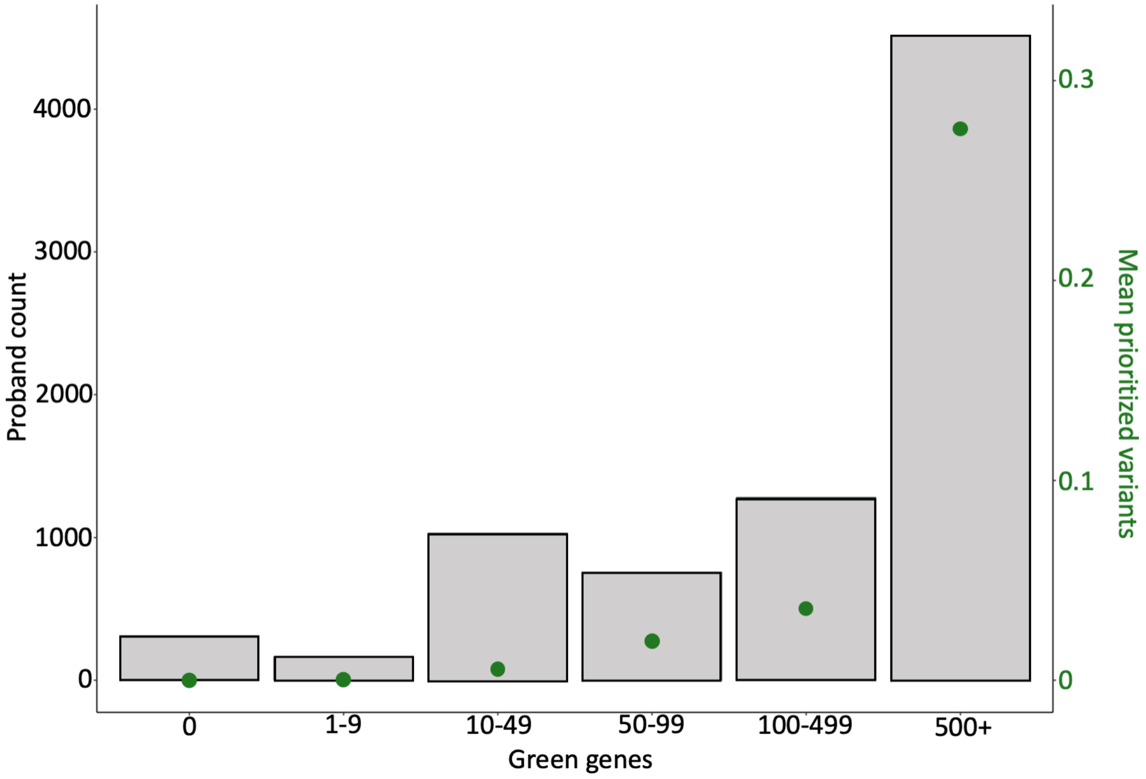

**Supplementary Figure 2**: The number of probands with different numbers of assigned green-dominant genes (grey bars) and mean prioritised variants from the *de novo* variant dataset (green dots) in 8,040 probands.

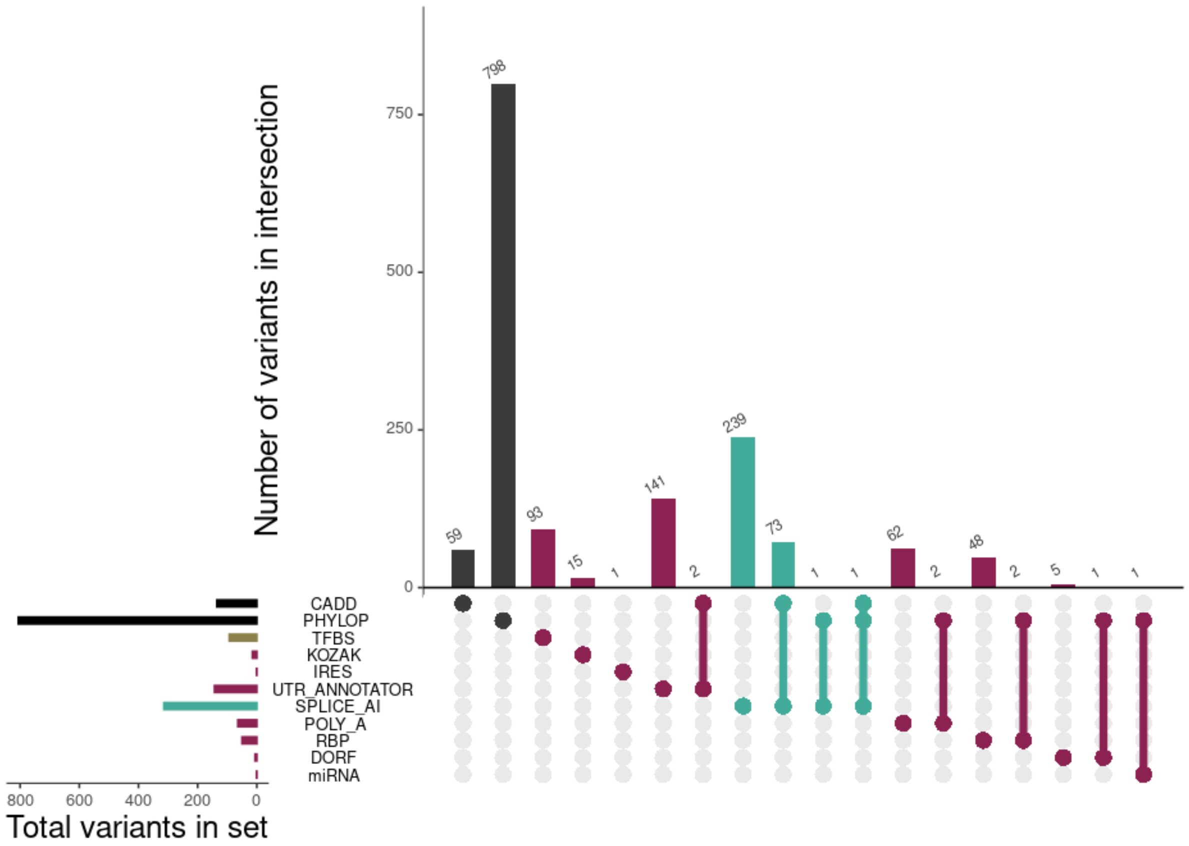

**Supplementary figure 3**: Annotated variants in the aggregated variant set used for burden testing. Universal variant annotations are shown in black, UTR specific variant annotations in raspberry, promoter specific variant annotations in mustard, and intronic / splicing variant annotations in teal. Vertical bars in the top panel denote the number of variants identified with specific region and variant annotations that are represented by the bar colour (regions), and in the upset plot  below (variant annotations). The total number of DNVs with each variant annotation is shown by the horizontal bars to the left of the upset.

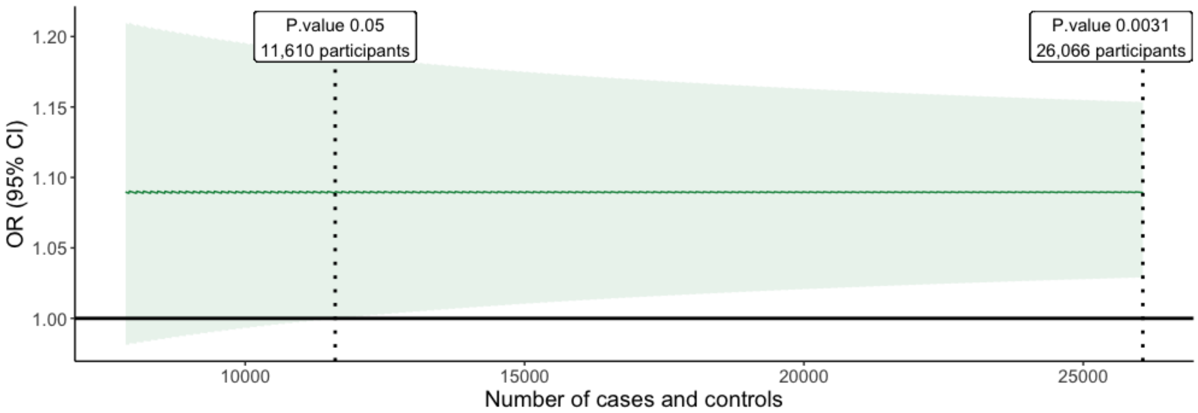

**Supplementary figure 4**: Estimated number of participants required to reach significance of *P<0.05* (left dotted line) and *P<0.0031* (right dotted line; Bonferroni adjusted threshold accounting for 16 tests) across all region and variant annotations. Odds ratio (OR) is shown in dark green, with 95% confidence interval shaded in light green. OR=1 is marked with a black solid line.
